# Better performance of deep learning pulmonary nodule detection using chest radiography with reference to computed tomography: data quality is matter

**DOI:** 10.1101/2023.02.09.23285621

**Authors:** Jae Yong Kim, Wi-Sun Ryu, Dongmin Kim, Eun Young Kim

## Abstract

**Background:** Labeling error may restrict radiography-based deep learning algorithms in screening lung cancer using chest radiography. Physicians also need precise location information for small nodules. We hypothesized that a deep learning approach using chest radiography data with pixel-level labels referencing computed tomography enhances nodule detection and localization compared to a data with only image-level labels.

**Methods:** National Institute Health dataset, chest radiograph-based labeling dataset, and AI-HUB dataset, computed tomography-based labeling dataset were used. As a deep learning algorithm, we employed Densenet with Squeeze-and-Excitation blocks. We constructed four models to examine whether labeling based on chest computed tomography versus chest X-ray and pixel-level labeling versus image-level labeling improves the performance of deep learning in nodule detection. Using two external datasets, models were evaluated and compared.

**Results:** Externally validated, the model trained with AI-HUB data (area under curve [AUC] 0.88 and 0.78) outperformed the model trained with NIH (AUC 0.71 and 0.73). In external datasets, the model trained with pixel-level AI-HUB data performed the best (AUC 0.91 and 0.86). In terms of nodule localization, the model trained with AI-HUB data annotated at the pixel level demonstrated dice coefficient greater than 0.60 across all validation datasets, outperforming models trained with image-level annotation data, whose dice coefficient ranged from 0.36-0.58.

**Conclusion:** Our findings imply that precise labeled data are required for constructing robust and reliable deep learning nodule detection models on chest radiograph. In addition, it is anticipated that the deep learning model trained with pixel-level data will provide nodule location information.

## Introduction

Pulmonary disorders are the top global causes of morbidity, mortality, and utilization of health services.^1^ Due to its accessibility, relative affordability, and widespread availability in outpatient clinics, chest radiography is the most widely performed diagnostic test in everyday medical practice.^2^ It is also widely used to screen lung cancer, the leading cause of cancer-related death for both men and women in the United States.^3^ Because missed pulmonary nodule or mass at chest radiography can result in delayed diagnoses and management for benign and malignant conditions, every image should be promptly reported by a radiologist. However, this is not always possible due to the high volume of work in many large healthcare facilities or the lack of experienced radiologists in less developed regions.^4-6^

Deep learning has shown a remarkable success in recent years.^7^ Using deep learning with convolutional neural networks in radiography has yielded excellent results in the diagnosis of numerous pulmonary disorders, such as pulmonary tuberculosis, pneumonia, and lung nodule.^8-11^ Prior research in deep learning for lung nodule detection were trained on large datasets, including National Institute of Health (NIH) Chest X-ray 14,^12^ CheXpert,^13^ and MIMIC-CXR.^14^ However, the performance of these algorithms are likely constrained by the dataset’s class imbalance and label noise resulting from disease label extraction process.^15, 16^ Incorrect labels hinder the generalization of predictive models and the validity of model evaluation during training and testing, respectively.^17^ Hence, label cleaning is therefore crucial to improve both model training and evaluation. In addition, in terms of pulmonary nodule detection, these algorithms offered limited information on the precise location, which is crucial due to the small and localized nature of pulmonary nodules.^18^ Consequently, high-quality data referenced to chest computed tomography (CT) with pixel-level label is desired in pulmonary nodule detection using deep learning algorithm.^19^

In the present study, we hypothesized that a deep learning algorithm utilizing chest radiography data with pixel-level labels referencing to chest CT improves nodule detection and localization performance when compared to a dataset containing only image-level labels.

## Methods

Our study was conducted with approval from the institutional review boards (IRB) of all participating centers (JLK Inc., and Gil medical center), and exemption from IRB review for AI-HUB datasets. The requirement for informed consent was waived.

### Datasets

Using two distinct datasets, we compared the nodule classification and localization performance between data with a noisy label and data with a clean label based on chest computed tomography. Randomly, the datasets were divided into train (70%), tune (10%), and validation (20%) sets. We utilized NIH Chest X-ray 14 dataset^12^ as big data with a noisy label. In Chest X-ray 14 dataset, a total of 112,120 frontal chest radiographs and their accompanying text reports were retrospectively retrieved from the PACS database of the NIH Clinical Center. As abnormal cases, we extracted 3,609 chest radiographs labeled only with nodule or mass. In addition, an equal number of normal cases were selected at random to match the number of abnormal cases. We used chest X-ray data from AI-HUB, a public dataset from the Republic of Korea, as a dataset with clean labels. The AI-HUB data set was retrospectively collected from a university hospital. The presence of a nodule on a chest radiograph was confirmed by experienced thoracic radiologists using chest CT as a reference standard. In addition, the location of the nodule was marked with a bounding box with referencing to CT. We created a pixel-level mask image for model training by drawing an ellipse within the bounding box. We used 3,177 out of 3,500 chest radiographs of nodules as abnormal cases, excluding low-quality images. In addition, 3,177 chest radiographs were randomly extracted from 10,000 normal chest X-ray images. In the AI-HUB dataset, duplicate patient images were not allowed.

We utilized two distinct chest radiograph datasets for external validation. One was collected retrospectively at the Gachon University Gil Medical Center (GMC). These chest radiographs included 246 cases of nodules and 440 cases of normal. A radiologist labeled them by referencing chest CT and annotated the nodule locations using pixel-level mask images. The other dataset was the VinBig dataset (VBD)^20^ made available to the public. It consisted of 15,000 chest radiographs collected in Vietnam. Three experienced radiologists annotated each chest radiograph’s lesion areas with a bounding box. We used as abnormal cases 83 chest radiographs annotated by two or more radiologists as having a nodule or mass in the same location.

### Deep convolutional neural network structure and development

From NIH and AI-HUB data, we randomly samples normal and nodule or mass images with a ratio of 1:1, taking class balance into account (Table 1). We built 4 models to investigate whether 1) labeling based on chest computed tomography versus chest X-ray and 2) pixel-level labeling versus image-level labeling improves deep learning’s performance in the nodule detection. The first model, the deep learning network pre-trained on ImageNet was transferred to NIH data. The second model, the network pre-trained on ImageNet was transferred to AI-HUB data with an image-level label. The third model, the network pre-trained on NIH data was transferred to AI-HUB data with an image-level label. For the fourth model, the network pre-trained on NIH data transferred to AI-HUB with a pixel-level label.

We built two neural networks, as depicted in Supplementary Fig 1, using Densenets with Squeeze-and-Excitation (SE) blocks and sub-networks. Using two convolution layers, the sub-network decreases the number of channels while preserving the width and height of the input feature maps. The network 1, which is employed for the classification task with image-level labeled data, is a standard Densenet with SE blocks. Network 1 analyzes the input chest x-ray and calculates the probability that a nodule is present, which ranges from 0 to 1. In order for the network to calculate loss between projected likelihood and a true label, the final global average pooling layer converts the output from the sub-network, which is 16 × 16 sized feature maps, to a float. Also, Network 1 provides a class activation map. Network 2 modified Network 1 to use a label at the pixel level. As a result, Network2 constructed a single-channel probability map from the output of the sub-network. We added an upsampling layer between Densenet and the sub-network, thereby using a two-fold upscaled feature map as the input for the sub-network, because a 16 × 16 sized feature map produced from Densenet with se-block is constrained to pinpoint the nodule position. The probability map shows the nodule’s location, and its maximum value denotes the probability that a nodule is present. In order to train Network2, classification and localization loss were summated. As the localization loss, we used the mean of the binary cross entropy at the pixel level. The probability map generated by the deep learning model and the 32×32 resized nodule mask from the ground truth were used to calculate the localization loss function.

In the training phase, we cropped an image around lung area to prevent background information of original image from interfering with network training.^21^ Then, we resized the cropped image to 512×512 and fed it as an input to the network. Also, for better performance and robustness, we applied contrast limited adaptive histogram equalization (CLAHE) and augmentation techniques such as horizontal flipping, shifting, scaling, rotating and shearing. We trained the network by using Adam optimizer with the batch size of 16. We set the initial learning rate of 0.0001 and then multiplied 0.1 at 25th, 37th epoch. Our experiments were based on Tensorflow v1.14.0 with Keras v2.3.0 and performed on an Intel® Xeon® Gold 5120 CPU @ 2.20GHz and 2 NVIDIA Tesla V100 GPUs.

### Quantification and statistical analysis

Each chest x-ray image was assigned one of four cases during the classification performance assessment by comparing the deep learning model’s prediction and the image-level ground truth label: true positive (TP), true negative (TN), false positive (FP), and false negative (FN). Specificity (Sp), sensitivity (Se), and area under (AUC) the receiver operating characteristic (ROC) curve were the three evaluation criteria we used. We utilized DeLong’s test for ROC curves to compare the AUC of different deep learning models.^22^

For localization performance assessment, we used the DICE coefficient score (DCS) between the class activation map or the heat map generated by deep learning models and ground truth mask annotated by thoracic radiologists. The metric is defined as follows:

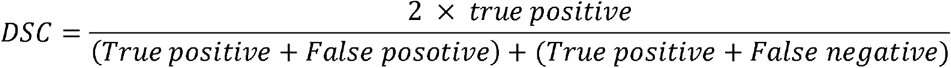

## Results

### Training and validation datasets

For model training, 7,218 radiographs (normal 3,609 and nodule 3,609) from the NIH dataset and 4,446 radiographs (normal 2,223 and nodule 2,223) from the AI-HUB dataset were used (Table 1). For the external validation, 686 cases (440 normal and 246 nodule) from GMC and 183 cases (100 normal and 83 nodule) from VBD were used.

### Comparison of model performance on nodule detection

Table 2, Figure 1, and Supplementary Fig 2 elaborate the performance summary of training algorithms on each dataset. Externally validated, the model trained with AI-HUB data (pretrained using ImageNet) outperformed the model trained with NIH data. In GMC and VBD datasets, AUCs for model trained with AI-HUB and NIH were 0.88 and 0.78 versus 0.71 and 0.73, respectively (p < 0.001). Notably, the model trained with AI-HUB data better performed on the dataset labeled with referencing to chest CT compared with dataset labeled with only chest radiograph (Figure 1C and Figure 1D). The model trained with NIH data outperformed the model trained with AI-HUB data only the internal validation data (Figure 1A). The model was trained with AI-HUB data after pretrain with NIH data improved performance only in VBD dataset (p < 0.001). Finally, the model trained with pixel-level AI-HUB data (pre-trained with NIH data) showed the best performance in both internal and external validation datasets. In GMC and VBD datasets, AUCs of the model trained with pixel-level data were 0.91 and 0.86, respectively.

**Figure 1.**
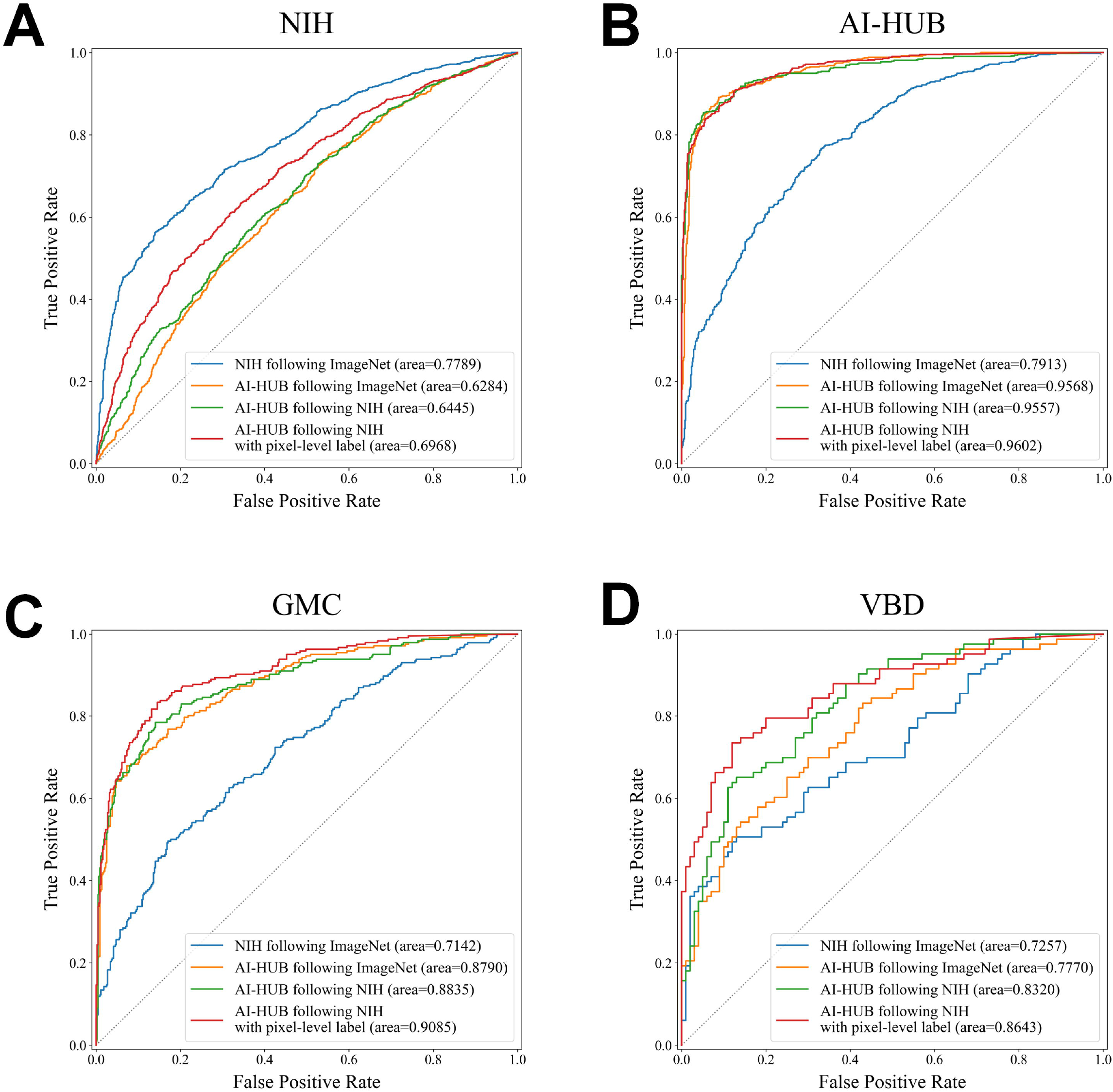

Figure 2 depicts the probability distributions of models for normal and nodule chest radiographs. Distributions of probability for normal and abnormal (nodule) images by the model trained with NIH data were considerably overlapping, particularly for external validation datasets, indicating poor discriminative performance. In contrast, the model trained on AI-HUB data effectively discriminated normal from abnormal chest X-ray data. The model trained with AI-HUB data demonstrated improved probability distribution even when validated against the NIH dataset, despite having lower AUC values than the model trained with NIH data.

**Figure 2.**
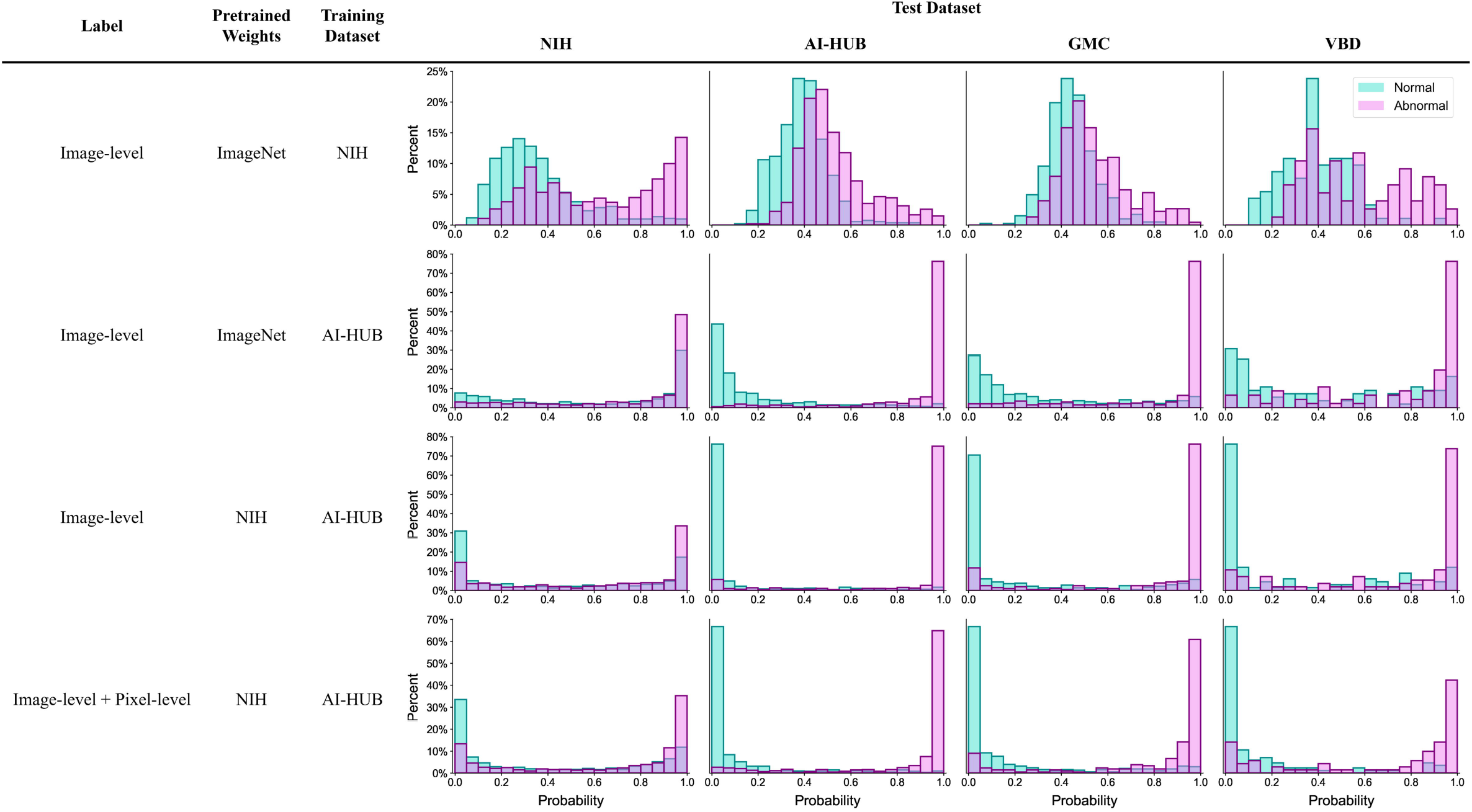

### Comparison of model performance on nodule localization

In external datasets with pixel-level annotations (GMC), we compared the trained models’ nodule localization performance. Models trained with image-level annotation data (the first, second, and third columns of Table 2) exhibited dice coefficient scores ranging from 0.36 to 0.58, which are comparable between models. However, the model trained with AI-HUB data annotated at the pixel level demonstrated dice coefficient values greater than 0.60 in all validation datasets. Figure 3 depicts representative radiographs demonstrating the nodule localization performance of the trained models.

**Figure 3.**
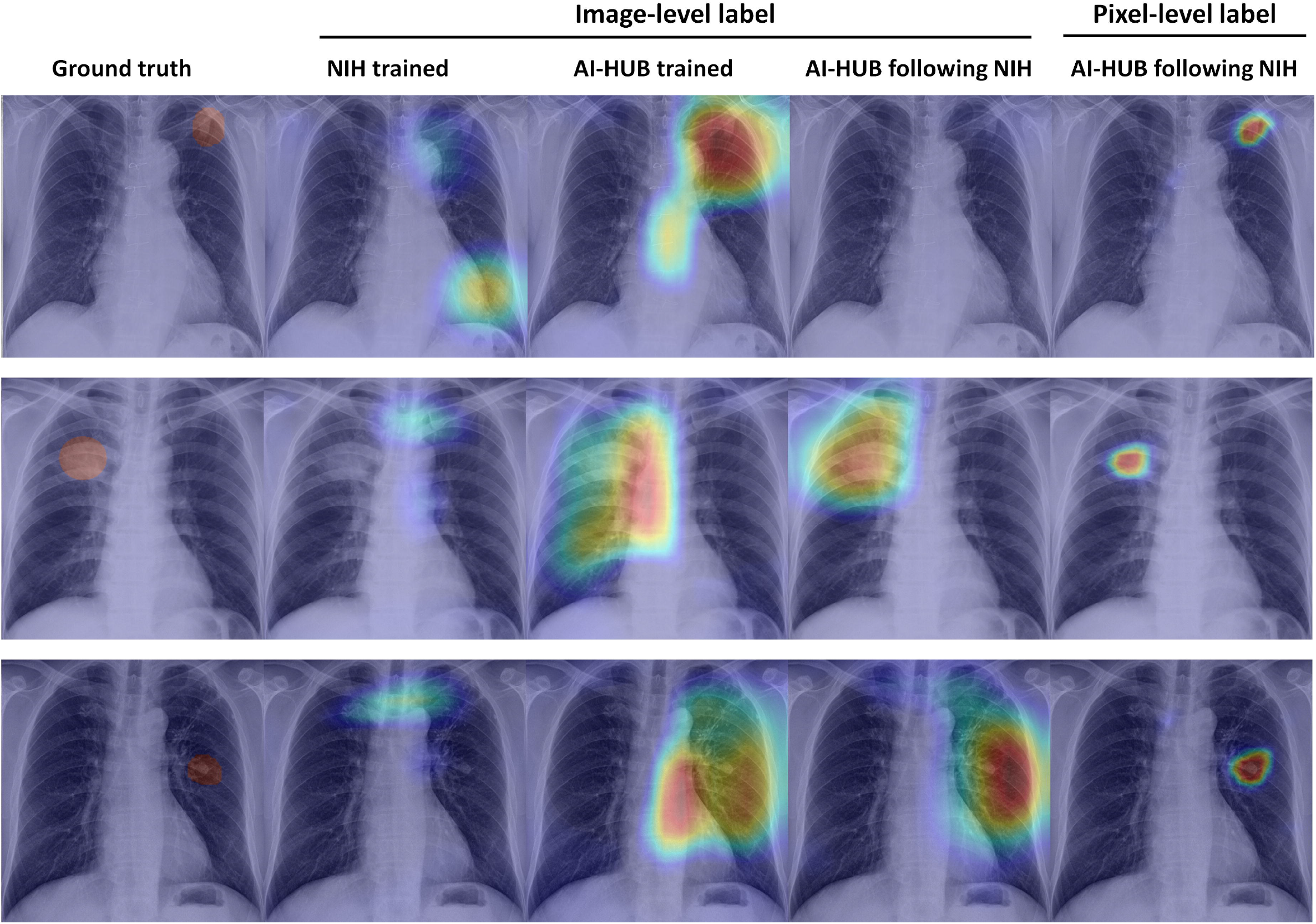

## Discussion

In the present study, we found that a deep learning model trained with chest radiographs data with a labeling referring to chest CT outperformed the model trained with chest radiographs data without exact labeling in a nodule detection. In addition, the performance of the deep learning model was enhanced by the addition of pixel-level annotations. On chest radiographs, the deep learning model trained with pixel-level label demonstrated more precise localization of nodules. Even in large datasets, precise labeling and detailed information are required to construct a precise deep learning model for the interpretation of medical images.

The NIH chest X-ray dataset is preprocessed using natural language processing; consequently, it tends to contain incorrect and uncertain labels.^16, 23^ Several reports on the effect of label noise on the classification of convolutional neural networks have demonstrated that CNNs are resistant to massive label noise.^24^ One study, however, argued that the deep learning–based computer-assisted diagnosis model is sensitive to label noise and that computer-assisted diagnosis with inaccurate labels is not credible.^25^ In addition, the study indicated that the deep learning performance is inversely related with a noisy label data ratio.^25^ In addition, the study revealed that the performance of deep learning is negatively correlated with the ratio of noisy label data.^26^ Our findings that the deep learning model trained with more precise label data outperformed the model trained with less precise label data are consistent with previous research. Taken together, given the consequences of misclassifying a medical image, the deep learning model in medical imaging requires precise data labeling.

In the present study, the model trained with accurate labels differentiated nodule x-rays from normal images more explicitly. Although deep learning in medical imaging has made tremendous strides, there are still a number of obstacles to overcome. The most significant disadvantage of deep learning in medical imaging is the ‘domain shift problem,’ in which the performance of the model degrades when applied to data from a different domain.^27^ Because the classification model classifies input data as normal or abnormal based on a threshold, the model producing highly overlapping probability distributions for normal and abnormal cases may be susceptible to the domain shift problem. In contrast, the model train with label based on CT showed a marked difference in distributions between nodule and normal chest x-rays, indicating the model’s robustness, as confirmed by two external validation datasets.

The class activation map generated by a deep learning classification model is a simple method for obtaining the discriminative image regions used by a neural network to identify a specific class in the image,^28^ which physicians frequently misinterpret as indicating the lesion area. Prior research has demonstrated that the class activation map often indicates disease-related regions rather than lesions per se.^29^ For instance, the class activation map produced by a deep learning model for detecting pneumothorax shows around the chest tube rather than pneumothorax itself.^30^ A study using the chest x-ray dataset for coronavirus disease 2019 (COVID-19) demonstrated conclusively that the classification deep learning algorithm uses “shortcuts” such as a laterality marker to distinguish between normal and abnormal cases.^31^ In the present study, the deep learning model was trained using two loss functions, nodule classification and localization, which is likely less susceptible to the “shortcuts” problem. Also, higher dice coefficient in the model trained with pixel-level data also buttressed this speculation. Due to the typically diminutive size of nodules on chest radiograph, providing a precise localization and accurate classification may be useful in daily clinical practice.

Our findings should be interpreted with caution. In the present study, the deep learning models were validated using two distinct external datasets. Due to the AI-HUB dataset and two external datasets collected from the Asian population, the observed performance difference in the study could be attributable, in part, to the ethnic variation of nodules.

The detection of lung nodules on a chest radiograph is crucial because lung nodules may indicate lung cancer. However, this is often difficult, and radiologists frequently miss lung nodules.^32^ Our findings indicate that accurate labeled data are required to build robust and reliable deep learning nodule detection models. In addition, it is expected that the deep learning model trained with pixel-level data will provide information on nodule location.

## Supporting information

tables and figures lagend

## Data Availability

The NIH chest radiographs that support the findings of this study are publicly
available at https://nihcc.app.box.com/v/ChestXray-NIHCC. And the VinBig dataset is publicly available at https://physionet.org/content/vindr-pcxr/1.0.0/.

https://nihcc.app.box.com/v/ChestXray-NIHCC

https://physionet.org/content/vindr-pcxr/1.0.0/

## DATA AVAILABILITY

The NIH chest radiographs that support the findings of this study are publicly available at https://nihcc.app.box.com/v/ChestXray-NIHCC. And the VinBig dataset is publicly available at https://physionet.org/content/vindr-pcxr/1.0.0/.

## CODE AVAILABILITY

The codes are not available.

## ACKNOWLEDGEMENTS

This research was supported by a grant from the Gachon University Gil Medical Center (Grant number: FRD2021-11).

## AUTHOR CONTRIBUTIONS

JY Kim, WS Ryu, and EY Kim devised the idea for the study. EY Kim collected data. JY Kim and WS Ryu analyzed and interpreted data. WS Ryu, and EY Kim provided overall supervision of the study and writing. JY Kim, WS Ryu, and EY Kim wrote the initial draft of the paper. JY Kim and WS Ryu verified the underlying data. All authors critically revised the manuscript and have seen and approved the final version to be published.

## References

1. Mortality GBD and Causes of Death C. Global, regional, and national life expectancy, all-cause mortality, and cause-specific mortality for 249 causes of death, 1980-2015: a systematic analysis for the Global Burden of Disease Study 2015. Lancet. 2016;388:1459–1544.

2. Brogdon BG, Kelsey CA and Moseley RD, Jr. Factors affecting perception of pulmonary lesions. Radiol Clin North Am. 1983;21:633–54.

3. Torre LA, Siegel RL and Jemal A. Lung Cancer Statistics. Adv Exp Med Biol. 2016;893:1–19.

4. Forrest JV and Friedman PJ. Radiologic errors in patients with lung cancer. West J Med. 1981;134:485–90.

5. Levin DC, Rao VM, Parker L and Frangos AJ. Analysis of radiologists’ imaging workload trends by place of service. J Am Coll Radiol. 2013;10:760–3.

6. Bhargavan M, Kaye AH, Forman HP and Sunshine JH. Workload of radiologists in United States in 2006-2007 and trends since 1991-1992. Radiology. 2009;252:458–67.

7. LeCun Y, Bengio Y and Hinton G. Deep learning. Nature. 2015;521:436–44.

8. Nam JG, Park S, Hwang EJ, Lee JH, Jin KN, Lim KY, Vu TH, Sohn JH, Hwang S, Goo JM, et al. Development and Validation of Deep Learning-based Automatic Detection Algorithm for Malignant Pulmonary Nodules on Chest Radiographs. Radiology. 2019;290:218–228.

9. Liu V, Clark MP, Mendoza M, Saket R, Gardner MN, Turk BJ and Escobar GJ. Automated identification of pneumonia in chest radiograph reports in critically ill patients. BMC Med Inform Decis Mak. 2013;13:90.

10. Hua KL, Hsu CH, Hidayati SC, Cheng WH and Chen YJ. Computer-aided classification of lung nodules on computed tomography images via deep learning technique. Onco Targets Ther. 2015;8:2015–22.

11. Lakhani P and Sundaram B. Deep Learning at Chest Radiography: Automated Classification of Pulmonary Tuberculosis by Using Convolutional Neural Networks. Radiology. 2017;284:574–582.

12. Wang X, Peng Y, Lu L, Lu Z and Summers RM. Tienet: Text-image embedding network for common thorax disease classification and reporting in chest x-rays. 2018:9049–9058.

13. Irvin J, Rajpurkar P, Ko M, Yu Y, Ciurea-Ilcus S, Chute C, Marklund H, Haghgoo B, Ball R and Shpanskaya K. Chexpert: A large chest radiograph dataset with uncertainty labels and expert comparison. 2019;33:590–597.

14. Johnson AE, Pollard TJ, Greenbaum NR, Lungren MP, Deng C-y, Peng Y, Lu Z, Mark RG, Berkowitz SJ and Horng S. MIMIC-CXR-JPG, a large publicly available database of labeled chest radiographs. arXiv preprint arXiv:190107042. 2019.

15. Rajpurkar P, Irvin J, Ball RL, Zhu K, Yang B, Mehta H, Duan T, Ding D, Bagul A, Langlotz CP, et al. Deep learning for chest radiograph diagnosis: A retrospective comparison of the CheXNeXt algorithm to practicing radiologists. PLoS Med. 2018;15:e1002686.

16. Oakden-Rayner L. Exploring Large-scale Public Medical Image Datasets. Acad Radiol. 2020;27:106–112.

17. Arpit D, Jastrzę bski S, Ballas N, Krueger D, Bengio E, Kanwal MS, Maharaj T, Fischer A, Courville A and Bengio Y. A closer look at memorization in deep networks. 2017:233–242.

18. Loverdos K, Fotiadis A, Kontogianni C, Iliopoulou M and Gaga M. Lung nodules: A comprehensive review on current approach and management. Ann Thorac Med. 2019;14:226–238.

19. Bernhardt M, Castro DC, Tanno R, Schwaighofer A, Tezcan KC, Monteiro M, Bannur S, Lungren MP, Nori A, Glocker B, et al. Active label cleaning for improved dataset quality under resource constraints. Nat Commun. 2022;13:1161.

20. Nguyen HQ, Lam K, Le LT, Pham HH, Tran DQ, Nguyen DB, L. DD, Pham CM, Tong HT and Dinh DH. VinDr-CXR: An open dataset of chest X-rays with radiologist’s annotations. Scientific Data. 2022;9:429.

21. Teixeira LO, Pereira RM, Bertolini D, Oliveira LS, Nanni L, Cavalcanti GD and Costa YM. Impact of lung segmentation on the diagnosis and explanation of COVID-19 in chest X-ray images. Sensors. 2021;21:7116.

22. DeLong ER, DeLong DM and Clarke-Pearson DL. Comparing the areas under two or more correlated receiver operating characteristic curves: a nonparametric approach. Biometrics. 1988;44:837–45.

23. Rajpurkar P, Irvin J, Zhu K, Yang B, Mehta H, Duan T, Ding D, Bagul A, Langlotz C and Shpanskaya K. Chexnet: Radiologist-level pneumonia detection on chest x-rays with deep learning. arXiv preprint arXiv:171105225. 2017.

24. Rolnick D, Veit A, Belongie S and Shavit N. Deep learning is robust to massive label noise. arXiv preprint arXiv:170510694. 2017.

25. Jang R, Kim N, Jang M, Lee KH, Lee SM, Lee KH, Noh HN and Seo JB. Assessment of the robustness of convolutional neural networks in labeling noise by using chest X-ray images from multiple centers. JMIR medical informatics. 2020;8:e18089.

26. Karimi D, Dou H, Warfield SK and Gholipour A. Deep learning with noisy labels: Exploring techniques and remedies in medical image analysis. Med Image Anal. 2020;65:101759.

27. Guan H and Liu M. Domain Adaptation for Medical Image Analysis: A Survey. IEEE Trans Biomed Eng. 2022;69:1173–1185.

28. Zhou B, Khosla A, Lapedriza A, Oliva A and Torralba A. Learning deep features for discriminative localization. Proceedings of the IEEE conference on computer vision and pattern recognition. 2016:2921–2929.

29. Huff DT, Weisman AJ and Jeraj R. Interpretation and visualization techniques for deep learning models in medical imaging. Phys Med Biol. 2021;66:04TR01.

30. Seah J, Tang C, Buchlak QD, Milne MR, Holt X, Ahmad H, Lambert J, Esmaili N, Oakden-Rayner L, Brotchie P, et al. Do comprehensive deep learning algorithms suffer from hidden stratification? A retrospective study on pneumothorax detection in chest radiography. BMJ Open. 2021;11:e053024.

31. DeGrave AJ, Janizek JD and Lee SI. AI for radiographic COVID-19 detection selects shortcuts over signal. medRxiv. 2020.

32. Gavelli G and Giampalma E. Sensitivity and specificity of chest x-ray screening for lung cancer. Cancer. 2000;89:2453–2456.

