## supplemental for "Better performance of deep learning pulmonary nodule detection using chest radiography with reference to computed tomography: data quality is matter"

Supplementary Fig 1. Deep learning network for training

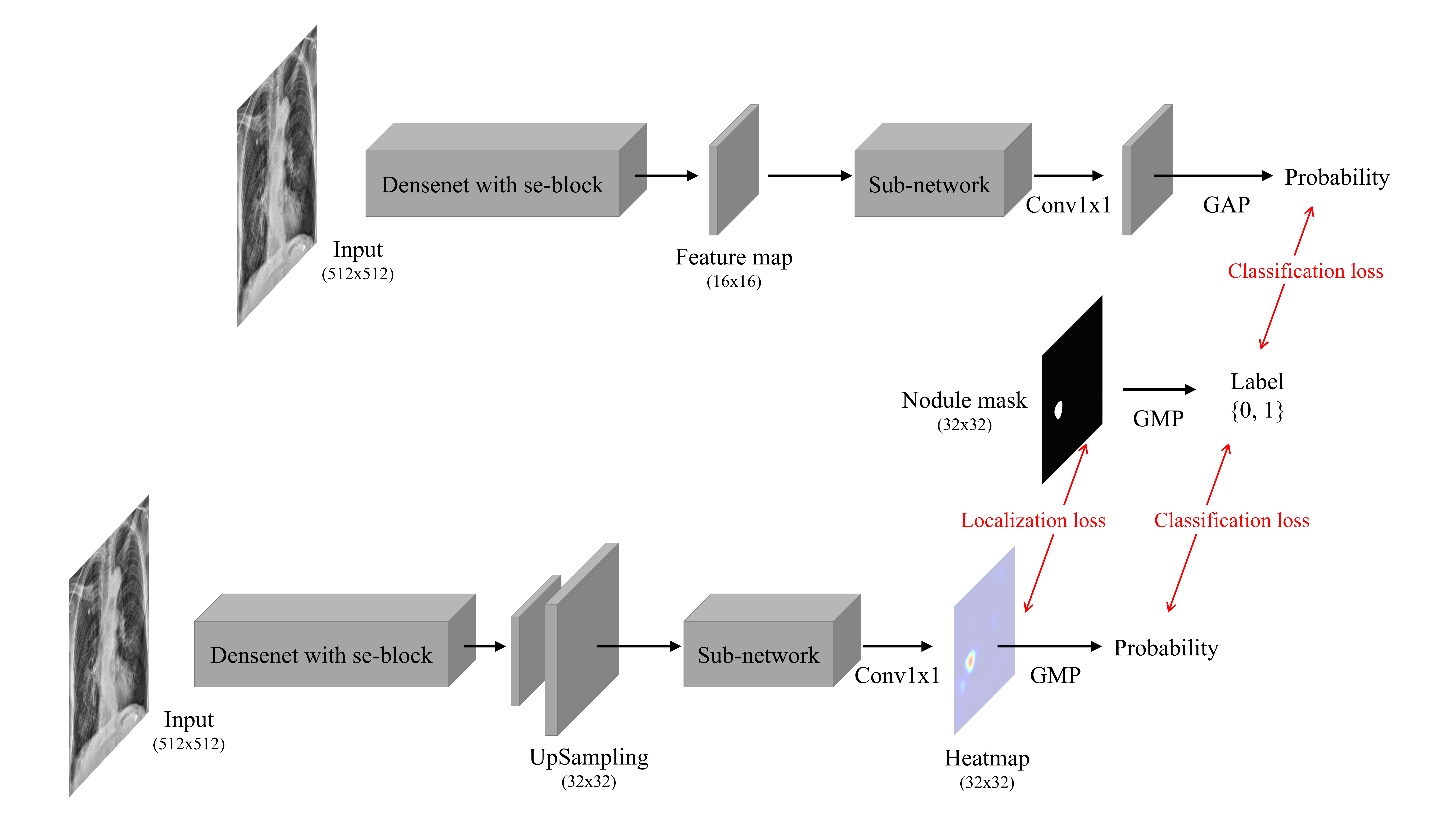

Supplementary Fig 2. Confusion matrix for four different test datasets

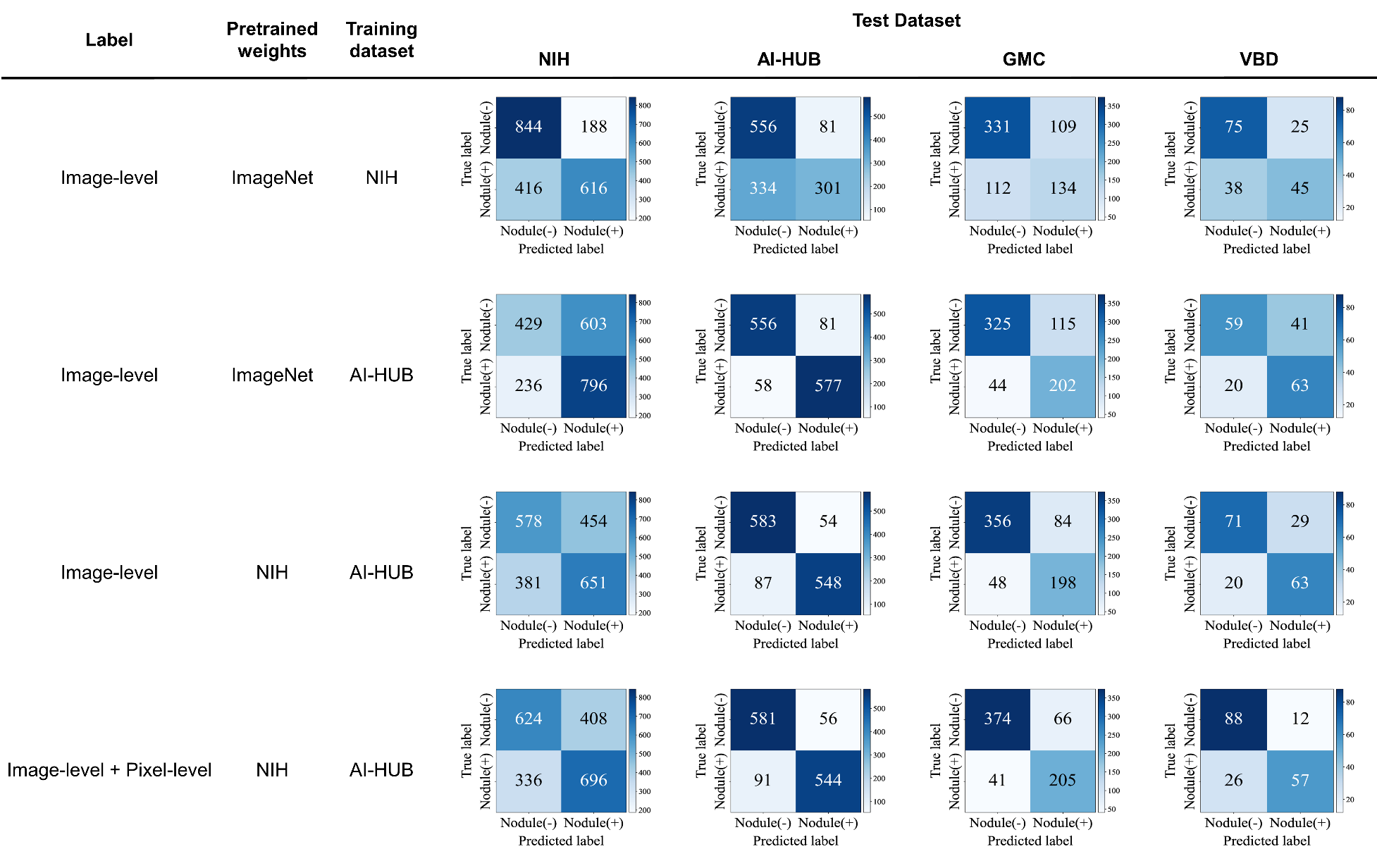
