## Supplementary material for "Better performance of deep learning pulmonary nodule detection using chest radiography with reference to computed tomography: data quality is matter": tables and figures lagend

**Table 1. Training, tuning and validation datasets**

|  | **Train** | | **Tune** | | **Validation** | | | |
| --- | --- | --- | --- | --- | --- | --- | --- | --- |
|  |  |  |  |  | **Internal** | | **External** | |
|  | **NIH** | **AI-HUB** | **NIH** | **AI-HUB** | **NIH** | **AI-HUB** | **GMC** | **VBD** |
| No. of chest radiographs | 7218 | 4446 | 1030 | 636 | 2064 | 1272 | 686 | 183 |
| No. of normal radiographs | 3609 | 2223 | 515 | 318 | 1032 | 636 | 440 | 100 |
| No. of nodule radiographs | 3609 | 2223 | 515 | 318 | 1032 | 636 | 246 | 83 |

The NIH and AI-HUB datasets were divided 7:1:2 into training, tuning, and validation data. NIH = National Institute of Health; GMC = Gachon medical center; VBD = VinBig data.

**Table 2. Performances of four different deep learning algorithms in four different dataset**

|  |  | | **Image level data** | | | | | | | | | | | **Pixel level data** | | | |
| --- | --- | --- | --- | --- | --- | --- | --- | --- | --- | --- | --- | --- | --- | --- | --- | --- | --- |
| **Validation data** | **ImageNet 🡪 NIH** | | | | | **ImageNet 🡪 AI-HUB** | | | | **ImageNet 🡪 NIH 🡪 AI-HUB** | | | | **ImageNet 🡪 NIH 🡪 AI-HUB** | | | |
|  | **AUC** | **Sp.** | | **Se.** | **DSC** | **AUC** | **Sp.** | **Se.** | **DSC** | **AUC** | **Sp.** | **Se.** | **DSC** | **AUC** | **Sp.** | **Se.** | **DSC** |
| NIH | 0.78 | 0.82 | | 0.60 | NA | 0.63 | 0.42 | 0.77 | NA | 0.64 | 0.56 | 0.63 | NA | 0.70 | 0.60 | 0.67 | NA |
| AI-HUB | 0.79 | 0.87 | | 0.47 | 0.48 | 0.96 | 0.87 | 0.91 | 0.49 | 0.96 | 0.92 | 0.86 | 0.51 | 0.96 | 0.91 | 0.86 | 0.65 |
| GMC | 0.71 | 0.75 | | 0.54 | 0.51 | 0.88 | 0.74 | 0.82 | 0.53 | 0.88 | 0.81 | 0.80 | 0.58 | 0.91 | 0.85 | 0.83 | 0.64 |
| VBD | 0.73 | 0.75 | | 0.54 | 0.46 | 0.78 | 0.59 | 0.76 | 0.36 | 0.83 | 0.71 | 0.76 | 0.42 | 0.86 | 0.88 | 0.69 | 0.60 |

AUC = Area under curve; Sp. = specificity; Se. = sensitivity; DSC = Dice similarity coefficient; NIH = National Institute of Health; GMC = Gachon medical center; VBD = VinBig data.

**Figure Legends**

**Figure 1. Receive operating characteristics curve of four models validated in internal and external datasets**

A. Validation using NIH data, B. Validation using AI-HUB data, C. Validation using GMC data, D. Validation using VBD data. The NIH and VBD datasets were labeled using chest radiographs. Data labeling for AI-HUB and GMC was based on chest computed tomography.

NIH = National Institute of Health; GMC = Gachon Gil Medical Center; VBD = VinBing data

**Figure 2. Probability distributions of models trained with different datasets for normal and nodule chest radiographs**

NIH = National Institute of Health; GMC = Gachon Gil Medical Center; VBD = VinBing data

**Figure 3. Representative radiographs, activation map, or heatmap derived from deep learning models**

The first column indicates a thoracic radiologist's ground truth mask. The second, third, and fourth columns, respectively, represent activation maps obtained from models trained using NIH data, AI-HUB data, and AI-HUB data following NIH data with image-level label. The fifth column displays a heatmap obtained from a model trained with AI-HUB using NIH data labeled at the pixel level.

NIH = National Institute of Health
